# Strategies to reduce the risk of SARS-CoV-2 re-introduction from international travellers

**DOI:** 10.1101/2020.07.24.20161281

**Authors:** Samuel Clifford, Billy J. Quilty, Timothy W. Russell, Yang Liu, Yung-Wai Desmond Chan, Carl A. B. Pearson, Rosalind M. Eggo, Akira Endo, CMMID COVID-19 Working Group, Stefan Flasche, W. John Edmunds

## Abstract

To mitigate SARS-CoV-2 transmission risks from international travellers, many countries currently use a combination of up to 14 days of self-quarantine on arrival and testing for active infection. We used a simulation model of air travellers arriving to the UK from the EU or the USA and the timing of their stages of infection to evaluate the ability of these strategies to reduce the risk of seeding community transmission. We find that a quarantine period of 8 days on arrival with a PCR test on day 7 (with a 1-day delay for test results) can reduce the number of infectious arrivals released into the community by a median 94% compared to a no quarantine, no test scenario. This reduction is similar to that achieved by a 14-day quarantine period (median 99% reduction). Shorter quarantine periods still can prevent a substantial amount of transmission; all strategies in which travellers spend at least 5 days (the mean incubation period) in quarantine and have at least one negative test before release are highly effective (e.g. a test on day 5 with release on day 6 results in a median 88% reduction in transmission potential). Without intervention, the current high prevalence in the US (40 per 10,000) results in a higher expected number of infectious arrivals per week (up to 23) compared to the EU (up to 12), despite an estimated 8 times lower volume of travel in July 2020.

Requiring a 14-day quarantine period likely results in less than 1 infectious traveller each entering the UK per week from the EU and the USA (97.5th percentile). We also find that on arrival the transmission risk is highest from pre-symptomatic travellers; quarantine policies will shift this risk increasingly towards asymptomatic infections if eventually-symptomatic individuals self-isolate after the onset of symptoms. As passenger numbers recover, strategies to reduce the risk of re-introduction should be evaluated in the context of domestic SARS-CoV-2 incidence, preparedness to manage new outbreaks, and the economic and psychological impacts of quarantine.

## Background

With the introduction of non-pharmaceutical interventions (NPIs) such as physical distancing measures, many countries around the world have managed to curb local SARS-CoV-2 transmission and reduce the incidence of COVID-19 to sporadic cases and localised outbreaks. Under these circumstances, limiting re-introduction of infections from other countries becomes increasingly important in order to prevent additional outbreaks and avoid overwhelming resource-intensive control efforts.

The current guideline in a number of countries is self-quarantine of new arrivals either at their home, with family or friends, or hotels or other temporary accommodation for 14 days (1). It is expected that by day 14 at least 95% of eventually symptomatic cases have become symptomatic (2). However, the median incubation period for SARS-CoV-2 is about 5 days (2) and, assuming that travellers are equally likely to travel at any point in that period, a 5-day quarantine on arrival should suffice to allow more than 50% of the infections to develop symptoms and be managed accordingly. Quarantine, either at home or at managed facilities, may lead to negative psychological effects stemming from social isolation (3,4) and financial stress (5). In addition, the continued application of COVID-19 related travel restrictions, including quarantine, is likely to significantly impact economies reliant on tourist and business travel. On 7 May 2020 the United Nations World Tourism Organisation estimated that up to 80% of the USD1.7tn global earnings from tourism in 2019 may be lost in 2020, along with 120m jobs (6). In an IATA survey, quarantine was cited as the primary reason for reluctance to travel (85%), along with fear of becoming infected (84%), with 17% of respondents unwilling to undergo quarantine (7). Therefore, the anticipated personal, social and economic costs of extended quarantine must be justified by the reduction in transmission risk.

As well as quarantine, some countries have introduced a requirement for travellers to undergo testing for SARS-CoV-2 infection with Reverse Transcription Polymerase Chain Reaction (RT-PCR, hereafter PCR). Such testing is, commonly, performed by taking nasopharyngeal or throat swabs (NTS) of individuals and analysing the resulting sample for the presence of SARS-CoV-2 RNA (8). PCR screening may be conducted prior to the flight and/or upon arrival to allow detection of infected travellers on entry. Since 3 June 2020, Singapore has required visitors from China to take a PCR test no greater than 48-hours before departure, with a certificate of their infection-free status required for entry, and an additional test upon arrival (9). A similar policy is in place in Hong Kong (10); travellers who test positive on arrival are transferred to hospital, while those who test negative enter a compulsory 14 day quarantine period at home or hotel, and in a managed quarantine facility if they returned from perceived high-risk areas (11). In some countries, the testing strategy is also used to allow shortened or no quarantine for those without a confirmed infection. For example, Japan allows for business travellers from designated low-risk countries such as Australia, New Zealand, Thailand and Vietnam to skip the 14-day quarantine period if they test negative upon arrival (12). However, testing every arriving traveller introduces logistical constraints that may limit the possible traveller volume; e.g. Japan only allows 250 arriving travellers per day as of early June 2020 (13). As such, understanding the effectiveness of such strategies to limit the risk of re-introduction of SARS-CoV-2 is of vital importance.

Here we investigate a number of strategies targeting arriving travellers, including a combination of quarantine and screening for active infection. We consider the effectiveness of each strategy in reducing the number of infectious travellers who enter into the community and their remaining infectious duration.

## Methods

The travel screening outcomes for travellers are as follows: (i) prevented from travelling due to either syndromic screening at the airport or a positive pre-flight PCR test; (ii) released after the mandatory isolation period following either a positive PCR test at entry or a follow-up positive PCR test after a negative one at entry; (iii) released after a second negative test during the isolation period; (iv) In the absence of post-entry testing, travellers will either be free to enter immediately or be released after the mandatory isolation period (Figure 1).

**Figure 1.**
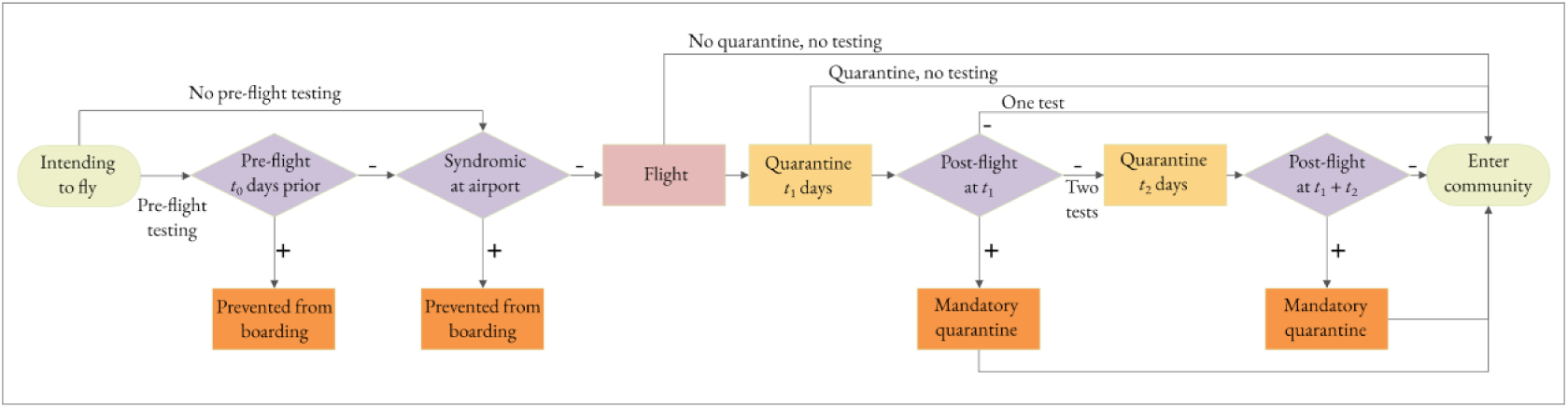
Possible traveller trajectories for the considered screening scenarios. Screening (purple diamonds) occurs pre-flight and/or post-flight and may include managed quarantine periods (yellow boxes). Travellers found to be infected pre-flight are prevented from boarding (orange boxes pre-flight); travellers found to be infected during managed quarantine are diverted to mandatory quarantine (orange boxes post-flight). Travellers enter the community after the required number of negative tests (regardless of infection status) or after meeting the requirements of the mandatory quarantine.

### Number of infected travellers

We simulated the number of infected air travellers intending to fly to a destination country in a given week based on the monthly volume of flights between origin and destination and the prevalence of COVID-19 in the origin country (Table 1). We used the United Kingdom of Great Britain and Northern Ireland (UK) as a case study for the destination country. We assumed that the inbound and outbound travel is balanced on average and halved the total number of monthly traveller movements, *n*, to estimate the number travelling into the UK. We sample the number of weekly arrivals, *W*, from a binomial distribution *W* ∼ *Bin*(*p* = 7/30, ⌈*n*/2⌉).

**Table 1:** Traveller movements in June 2019 and year on year change for May 2020 compared to May 2019 between UK airports, and airports in the European Union (EU) and United States of America (USA). Source: Civil Aviation Authority Tables 10.1 and 12.1 for July 2019 (14), May 2019 (14) and May 2020 (15).

|  | EU | USA | Source |
| --- | --- | --- | --- |
| Total traveller volume July 2019 | 18,186,680 | 2,249,856 | (14) |
| Year-on-year change for April and May 2020 compared to April and May 2019, % | -99% | -99% | (15)<br>EU: Table 10.1<br>USA: Table 12.1 |
| Calculated total traveller volume July 2020 using May year-on-year change, $n$ | 181,187 | 22,499 | (14,15)<br>EU: Table 10.1<br>USA: Table 12.1 |
| Duration of typical flight (hours) | 2 | 8 | Assumed |
| Prevalence of SARS-CoV-2 on 20 July 2020 | 2.8 per 10,000 | 40.0 per 10,000 | (16) |
| Number of infected individuals intending to travel in a given week. Median and 95% interval from 1000 simulations. | Symptomatic:<br>4 (1, 10)<br>Asymptomatic:<br>1 (0, 5) | Symptomatic:<br>8 (2, 21)<br>Asymptomatic:<br>2 (0, 10) | Proportion<br>asymptomatic derived<br>from (17) |

The time of each intending traveller’s flight was sampled uniformly between the time of their exposure to SARS-CoV-2 and their time of recovery. We assume that for the simulated travel scenarios, 70% of travellers who were symptomatic at their intended departure time were either prevented from travelling or chose not to travel (based on results from Figure 2 of Gostic et al. (2020) (18)). We modelled international travellers coming either from the European Union (EU) or the United States of America (USA), using publicly-available Civil Aviation Authority data for April and May 2020, which indicates that traveller volume was approximately 99% lower compared to the same period in 2019 (Table 1). The traveller volumes in July 2020 are therefore assumed to be approximately 1% of those in July 2019. Estimates of current COVID-19 infection prevalence were derived from reported cases and death time series data while adjusting for reporting delays and underreporting based on case-fatality ratio estimates (16,19). EU-wide prevalence was calculated as a population-weighted mean of available country-level estimates of the non-UK EU countries (except Malta, for which a prevalence estimate was not available). We assumed that 3-55% of infected intending travellers were asymptomatic (17).

**Figure 2.**
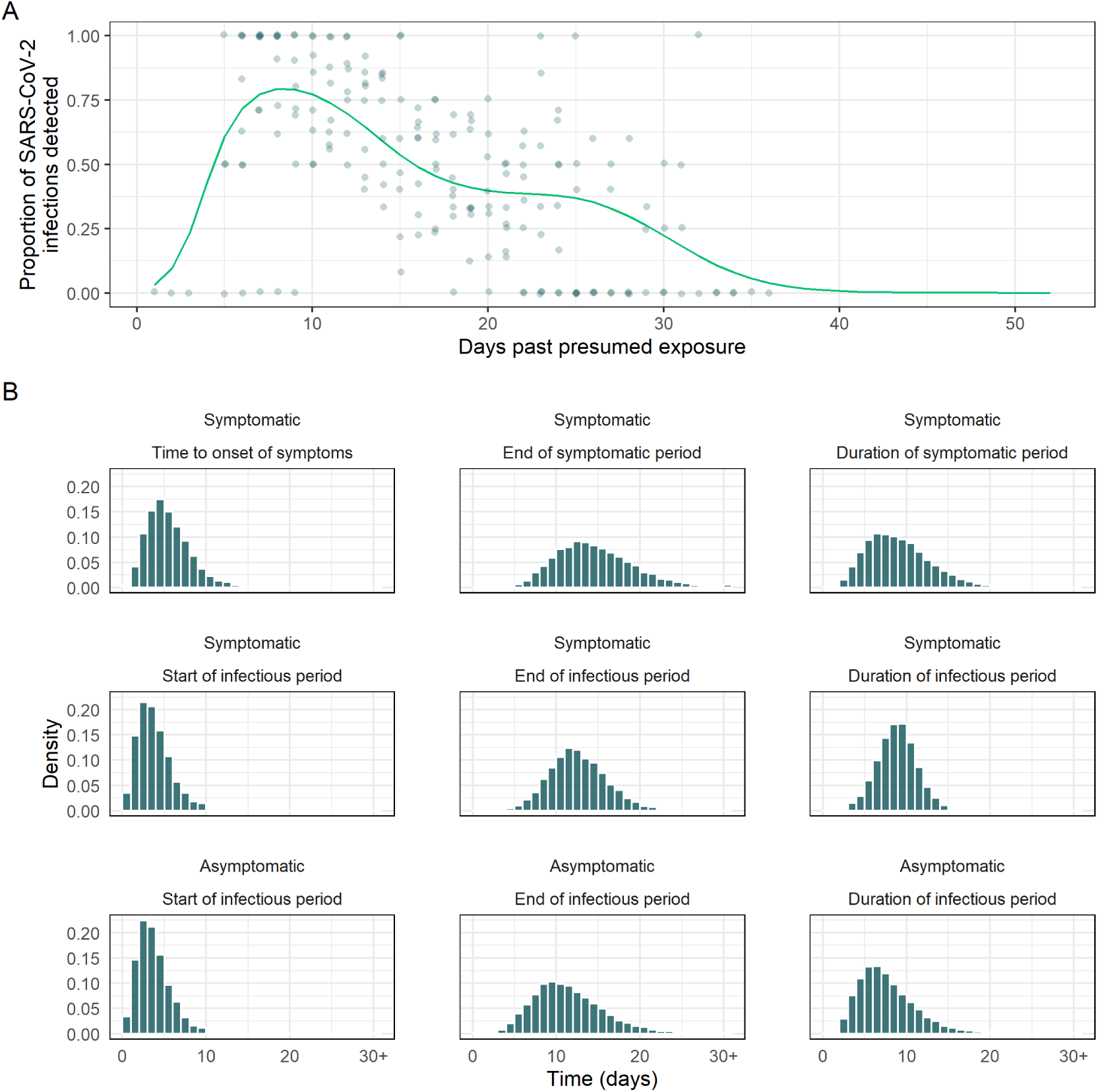
A. Traveller PCR sensitivity curves, obtained by fitting a Binomial GAM to the data collated in Kucirka et al. (2020) (25) The mean fit is used as the time-varying sensitivity function, *P*(*t*), and hence no uncertainty is shown in the figure. B. Distributions of times to clinically relevant events, namely time from exposure to start and end, and duration, of symptoms for symptomatic infections, and infectiousness for both symptomatic and asymptomatic infections. Times greater than 30 days are collapsed to a single “30+” bin.

We assume that the observed weekly travel volume, here, *W*, is those who have not been screened out or self-selected out based on onset of symptoms, i.e. the sum of the number of uninfected, asymptomatic, and those ever-symptomatic travellers not currently symptomatic. The total number of intending travellers, *W*′, is *W*, plus those who do not travel, *δW*. We calculate *W*′ as follows. First, sample *W* ∼ *Bin*(*p* = 7/30, ⌈*n*/2⌉). For *α*, the proportion of infections which are asymptomatic, *π*, the prevalence at the travel origin, *ξ*, the proportion of ever-symptomatic cases who are symptomatic at intended time of departure, and *ρ*, the proportion of currently symptomatic travellers prevented from boarding, *δW* is distributed according to a negative binomial distribution with size *W* and *p* = 1 − *π*(1 − *α*)*ρξ. ξ* is estimated by sampling a large number of ever-symptomatic travellers, along with flight departure times and symptomatic periods and determining which proportion are symptomatic at time of intended departure.

The number of uninfected travellers, *S*, is then *S* ∼ *Bin*(1 − *π, W* + *δW*); the number of asymptomatic infected travellers is *I*_*a*_ ∼ *Bin*(*α, W* + *δW* − *S*); the number of travellers symptomatic at time of departure is *I*_*s*_ ∼ *Bin*(*ξ, W* + *δW* − *S* − *I*_*a*_) and the number of ever-symptomatic travellers who are permitted to travel is therefore *W* + *δW* − *S* − *I*_*a*_ − *I*_*s*_ and is composed of those who are not yet symptomatic, those who are post-symptomatic, and those who are symptomatic but not detected by syndromic screening.

### Risk mitigation strategies

We considered several risk mitigation scenarios representing “low”, “moderate” and “high” levels of stringency, as well as that of a mandatory 14-day quarantine period (“maximum”) (Table 2, Figure 1). “High” was further stratified into multiple rounds of PCR testing: pre-flight, post-flight, and follow-up.

**Table 2.**
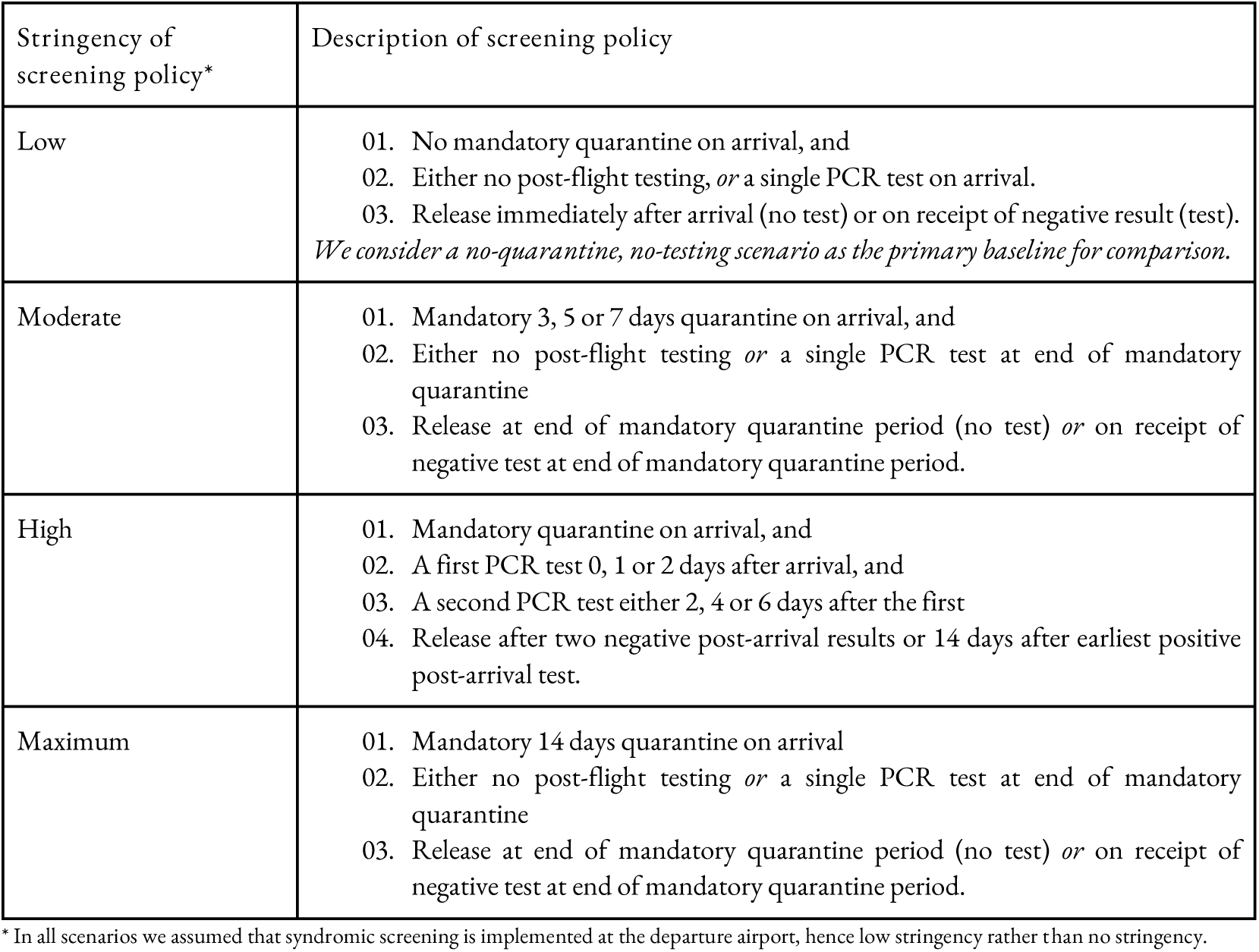
Strategies for risk mitigation. Where one of the described lines contains “or”, we consider all combinations contained within. For all levels of stringency we consider scenarios with the following pre-flight PCR policies: no pre-flight testing, pre-flight testing within 1 day of departure, within 4 days of departure, or within 1 week of departure.

| Stringency of screening policy* | Description of screening policy |
| --- | --- |
| Low | 01. No mandatory quarantine on arrival, and<br>02. Either no post-flight testing, *or* a single PCR test on arrival.<br>03. Release immediately after arrival (no test) or on receipt of negative result (test).<br>*We consider a no-quarantine, no-testing scenario as the primary baseline for comparison.* |
| Moderate | 01. Mandatory 3, 5 or 7 days quarantine on arrival, and<br>02. Either no post-flight testing *or* a single PCR test at end of mandatory quarantine<br>03. Release at end of mandatory quarantine period (no test) *or* on receipt of negative test at end of mandatory quarantine period. |
| High | 01. Mandatory quarantine on arrival, and<br>02. A first PCR test 0, 1 or 2 days after arrival, and<br>03. A second PCR test either 2, 4 or 6 days after the first<br>04. Release after two negative post-arrival results or 14 days after earliest positive post-arrival test. |
| Maximum | 01. Mandatory 14 days quarantine on arrival<br>02. Either no post-flight testing *or* a single PCR test at end of mandatory quarantine<br>03. Release at end of mandatory quarantine period (no test) *or* on receipt of negative test at end of mandatory quarantine period. |

\* In all scenarios we assumed that syndromic screening is implemented at the departure airport, hence low stringency rather than no stringency.

We assume that syndromic screening is performed prior to departure which may consist of thermal scanning and/or monitoring of symptoms such as cough and fever (20). Given the awareness of the pandemic and guidance issued on travelling while ill, we assume in all scenarios that 70% of currently-symptomatic travellers do not fly. This value, effectively a sensitivity of pre-flight syndromic screening for eventually symptomatic infections (Table 1), is derived from the post-incubation model results in Figure 2 of (18,21), adjusting the asymptomatic fraction to 0%.

Pre-flight PCR testing is required by some countries and airlines. Travellers must obtain a negative test result in a specific window prior to the flight’s departure. The IATA recommends testing within 24 hours before departure (22) but some countries require testing within the last seven days pre-flight (23). We consider pre-travel PCR at these two extremes and include a 4-day pre-flight test in order to provide a midpoint that demonstrates the transition between these extremes.

Travellers are subjected to a quarantine which lasts either zero (low stringency), 3, 5, or 7 (moderate stringency), or 14 (high and maximum stringency) days (Table 2). In all but the high level of stringency, travellers may also be tested on the final day of their quarantine (in the low stringency setting, this effectively enforces a one-day quarantine). At maximum stringency, the 14-day quarantine period aims to ensure that even a traveller who was infected just before or during the flight would likely spend their whole infectious period in quarantine and thereby not infect others. The moderately stringent strategy, on the other hand, aims to ensure that travellers spend a sufficient amount of time in quarantine to allow for the development of symptoms and probability of a positive PCR test leading to isolation for those infected. These strategies would, however, risk that some asymptomatically infected travellers (that is, infected travellers who will never display symptoms) will enter the community before the end of their infectious period. For the high stringency scenario, travellers are assumed to pass through multiple stages of PCR testing; if they test positive in either the immediate post-flight or follow-up testing, they will enter a mandatory isolation period, with positive pre-flight tests preventing an individual from flying altogether (we assume such travellers will defer their travel until they are cleared to fly). Travellers in the high stringency scenario will be cleared to leave quarantine upon receiving 2 negative tests during their post-arrival quarantine period, leaving quarantine the day after their final test to account for test delays of up to one day (24).

### Detection model

The time-varying PCR sensitivity function is the probability of an infected traveller testing positive by nasopharyngeal or throat swab (NTS) PCR on a given day, *P*(*t*). This is modelled as a function of the time since their exposure (Figure 1, Kucirka et al. 2020 (25)) fitting a Generalised Additive Model (GAM), with a Binomial likelihood and penalised B-spline basis (P-spline) (26), to the data collected by Kucirka et al. (2020) (25). We shift the observations, as they have, by an incubation period of 5 days (27), and augment by a pseudo-negative test on day 0 for each of the constituent data sets. We assumed that test specificity is 100% (28). We assumed that the probability of detecting an asymptomatic infection through PCR testing is 0.62 times that of a symptomatic individual’s infection, as reported by Chau et al. for NTS samples collected from quarantining travellers (29) (Table 3). The proportion of asymptomatic simulated travellers was derived by quantile matching the prediction interval of Buitrago-Garcia et al. (2020) as a Beta distribution with 95% interval (0.03, 0.55) (17).

**Table 3.**
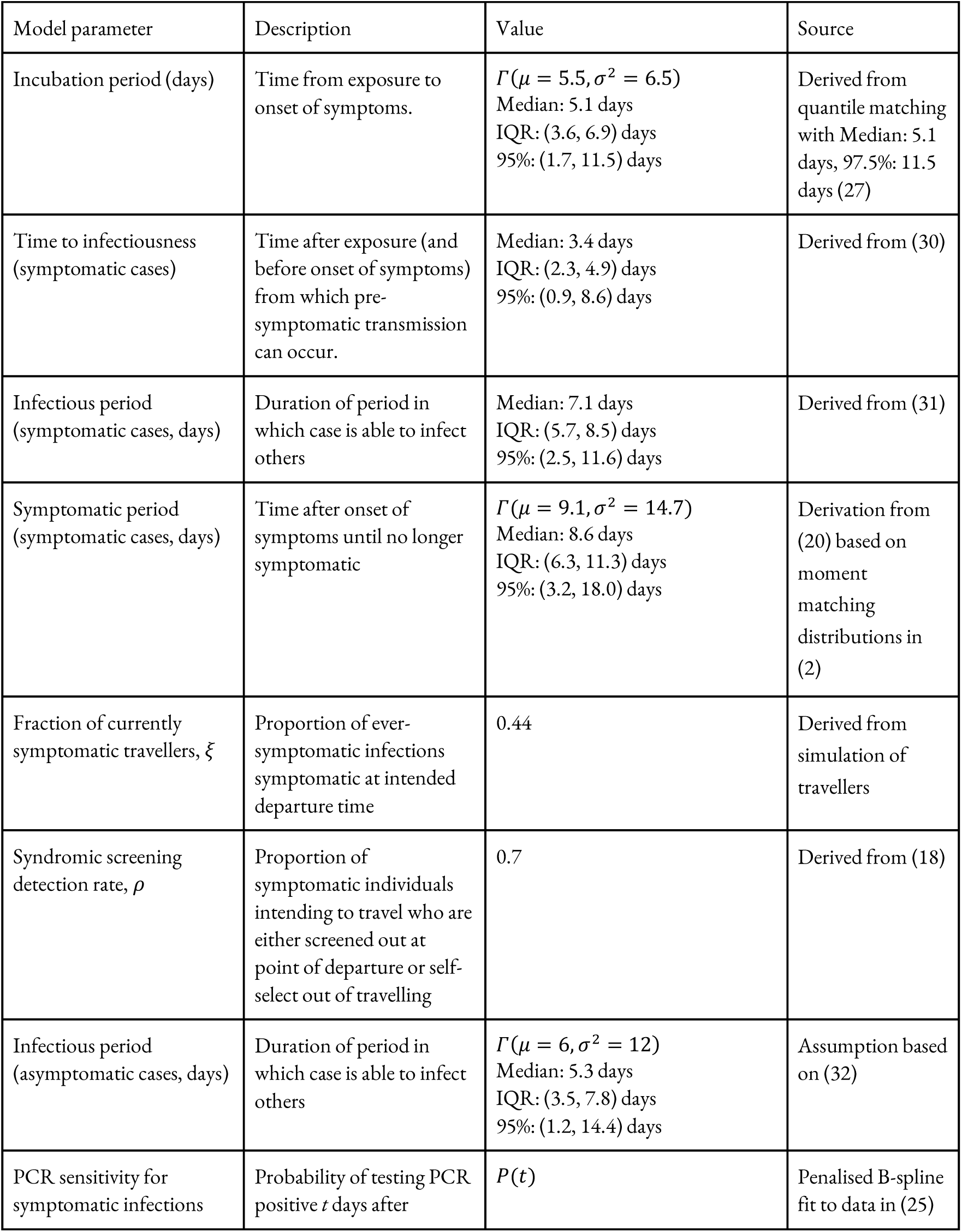
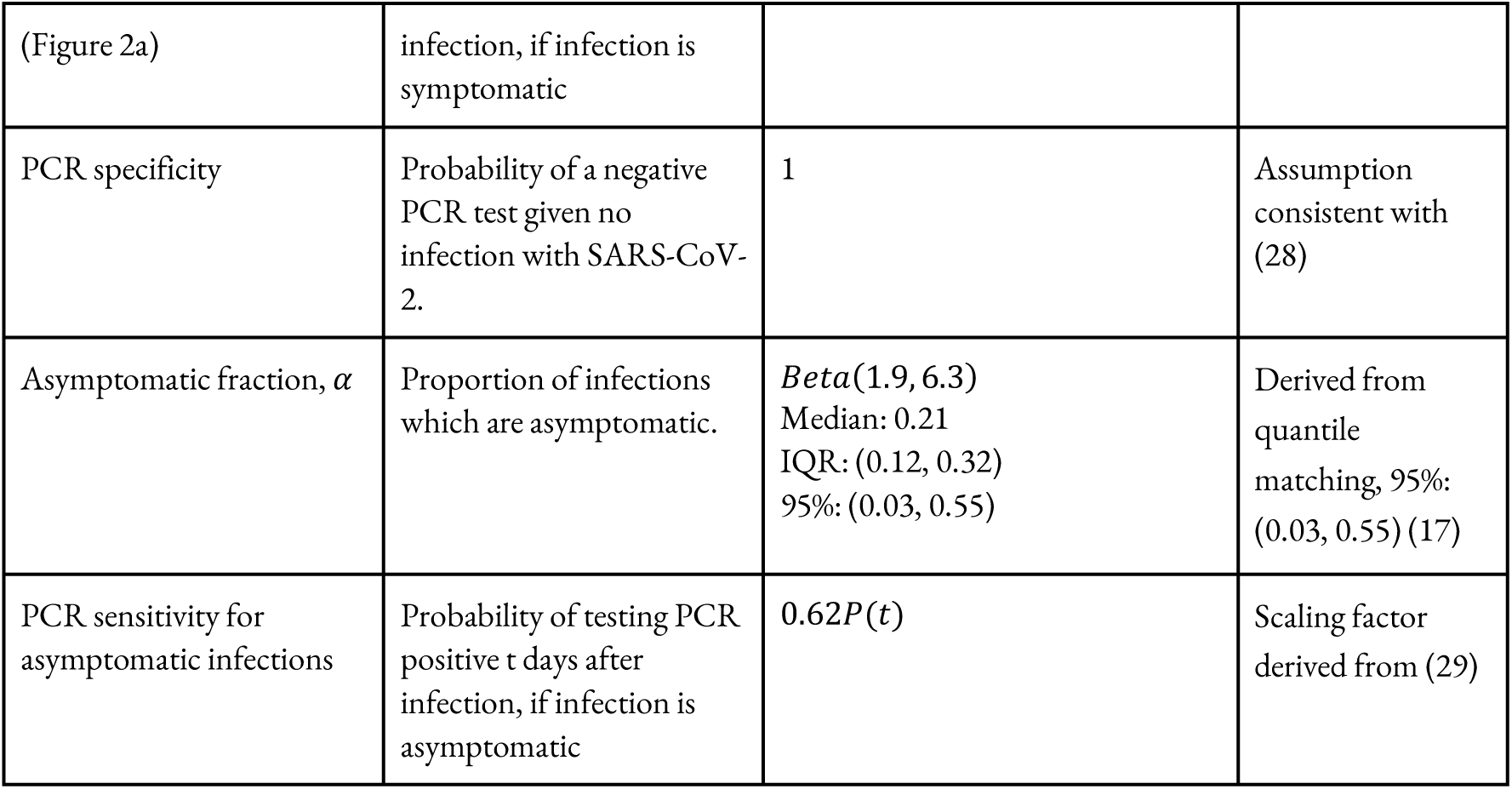
Values of parameters in simulation of travellers’ infection histories and PCR testing. Gamma distributions are parameterised in terms of a mean and variance, *Γ*(*μ, σ*^2^), and these are converted to shape and rate parameters via moment matching. Where quantiles are given but no distribution described, the parameter is derived from other distributions in the table and has no closed-form.

| Model parameter | Description | Value | Source |
| --- | --- | --- | --- |
| Incubation period (days) | Time from exposure to onset of symptoms. | $\Gamma(\mu = 5.5, \sigma^2 = 6.5)$<br>Median: 5.1 days<br>IQR: (3.6, 6.9) days<br>95%: (1.7, 11.5) days | Derived from quantile matching with Median: 5.1 days, 97.5%: 11.5 days (27) |
| Time to infectiousness (symptomatic cases) | Time after exposure (and before onset of symptoms) from which pre-symptomatic transmission can occur. | Median: 3.4 days<br>IQR: (2.3, 4.9) days<br>95%: (0.9, 8.6) days | Derived from (30) |
| Infectious period (symptomatic cases, days) | Duration of period in which case is able to infect others | Median: 7.1 days<br>IQR: (5.7, 8.5) days<br>95%: (2.5, 11.6) days | Derived from (31) |
| Symptomatic period (symptomatic cases, days) | Time after onset of symptoms until no longer symptomatic | $\Gamma(\mu = 9.1, \sigma^2 = 14.7)$<br>Median: 8.6 days<br>IQR: (6.3, 11.3) days<br>95%: (3.2, 18.0) days | Derivation from (20) based on moment matching distributions in (2) |
| Fraction of currently symptomatic travellers, $\xi$ | Proportion of ever-symptomatic infections symptomatic at intended departure time | 0.44 | Derived from simulation of travellers |
| Syndromic screening detection rate, $\rho$ | Proportion of symptomatic individuals intending to travel who are either screened out at point of departure or self-select out of travelling | 0.7 | Derived from (18) |
| Infectious period (asymptomatic cases, days) | Duration of period in which case is able to infect others | $\Gamma(\mu = 6, \sigma^2 = 12)$<br>Median: 5.3 days<br>IQR: (3.5, 7.8) days<br>95%: (1.2, 14.4) days | Assumption based on (32) |
| PCR sensitivity for symptomatic infections | Probability of testing PCR positive $t$ days after | $P(t)$ | Penalised B-spline fit to data in (25) |
| (Figure 2a) | infection, if infection is symptomatic |  |  |
| PCR specificity | Probability of a negative PCR test given no infection with SARS-CoV-2. | 1 | Assumption consistent with (28) |
| Asymptomatic fraction, $\alpha$ | Proportion of infections which are asymptomatic. | $Beta(1.9, 6.3)$<br>Median: 0.21<br>IQR: (0.12, 0.32)<br>95%: (0.03, 0.55) | Derived from quantile matching, 95%: (0.03, 0.55) (17) |
| PCR sensitivity for asymptomatic infections | Probability of testing PCR positive $t$ days after infection, if infection is asymptomatic | $0.62P(t)$ | Scaling factor derived from (29) |

Times to onset of symptoms, relevant for syndromic screening at point of departure, were sampled from Lauer et al. (2020) (27). We assume the same latent period for both symptomatic and asymptomatic cases (33), obtaining the parameters of the distribution by matching the median and 97.5% quantile of (27). Asymptomatic cases were assumed to have a mean of 6 days of infectiousness and a variance of 12 days, in line with the recommendations of Byrne et al. (2020) who discuss the difficulty in accurately characterising this period (32).

According to He et al. (2020) infectiousness of symptomatic cases begins 2.3 days (95%: (0.8, 3.0) days) prior to the onset of symptoms (30,34). We sampled this pre-symptomatic infectious period duration to derive the time from exposure to infectiousness by matching the quantiles of the distribution of time to onset of symptoms to the quantiles of the distribution of infectiousness lead times for each traveller, preserving order, ensuring that no time to infectiousness occurs before exposure. The duration of the infectious period for symptomatic cases was derived from the data of Wölfel et al. (2020) (31) by fitting a Binomial GAM with P-splines to determine the probability of no longer being infectious as a function of days since onset of symptoms. The time to non-infectiousness is sampled from the fitted GAM, which has range (0,1), by the inverse transform method (35).

The symptomatic period (defined by symptoms, sampled from a distribution derived by the authors in earlier work (20)) may differ from the infectious period (defined by viral load) and so we assume that the combination of syndromic screening and PCR testing will be more effective than either alone. Some travellers will be PCR-positive beyond their infectious period (29). Those testing positive on day 14 of the maximum stringency setting are quarantined for an additional 14 days. In line with current UK government guidance (1). Travellers who become symptomatic during their quarantine period must meet all of the following conditions prior to their release: they must no longer display symptoms; it must be at least seven days since the onset of symptoms; and they must have been in quarantine for at least 14 days. Persons later in their infection than the sum of the sampled (derived) exposure to infectiousness and duration of infectiousness were considered non-infectious and, hence, recovered, although they may still test positive under PCR (29,31).

We are therefore primarily interested in the number of individuals still in their infectious or pre-infectious period who are free to enter the UK in each of the scenarios, and report this quantity for the time spent in quarantine in each scenario.

In addition, we calculate the number of remaining infectious days for each infected traveller after their release from quarantine, reflecting the potential for onward transmission in the community. We report both the incidence per 10,000 and estimated weekly travellers based on travel volumes, with 1000 bootstraps replications each to generate medians and 95% and 50% uncertainty intervals. In addition, we calculate rate ratios in each screening scenario for the number of infectious persons released and infectious days remaining compared to the low scenario with syndromic screening, no quarantine, and no PCR testing (Figure 4). Rate ratios were calculated with 100,000 travellers per simulation to avoid small number biases and bootstrapped 500 times to generate medians and 95% and 50% uncertainty intervals. In the Supplementary Appendix we also include the total number of infectious person-days remaining (days remaining summed across still-infectious arrivals released).

All analysis was conducted in *R* version 4.0.2 (36); a link to the project Github repository can be found in the Supplementary Appendix.

## Results

Based on the prevalence of COVID-19 in the respective countries on 20 July 2020, we expect that the proportion of travellers who enter the UK infectious is substantially higher for flights originating in the USA than for those originating in the EU (Figure S1). However, as the prevalence of COVID-19 in the USA was approximately 14 times that in the EU in July 2020 and travel volumes were approximately 8 times lower than those in the EU, we expect approximately half the number of infectious arrivals from the EU than from the USA (Table 1). As a baseline for comparison, we use the lowest stringency scenario considered: 70% of currently symptomatic travellers are prevented from boarding, but no quarantine or testing is conducted. In this scenario, between 2 and 12 (EU), and 3 and 24 (USA) infectious travellers would enter the community (Figure 3A, low, no testing). By introducing a mandatory quarantine period of 7 days, this can be reduced to 0 to 3 infectious persons per week from the EU and 0 to 4 from the USA (Figure 3A, Mod.), preventing approximately 80% of travellers from entering the community while being infectious (Rate Ratios, median and 95% UI: EU: 0.18 (0.00, 0.42), USA: 0.18 (0.10, 0.27)) (Figure 4A). A mandatory quarantine period of 14 days resulted in 0 to 1 infectious entries per week each from the EU and USA (Figure 3A, Max.), an almost completely effective reduction (RR: EU: 0.00 (0.00, 0.01), USA: 0.01 (0.00, 0.04)) (Figure 4A).

**Figure 3.**
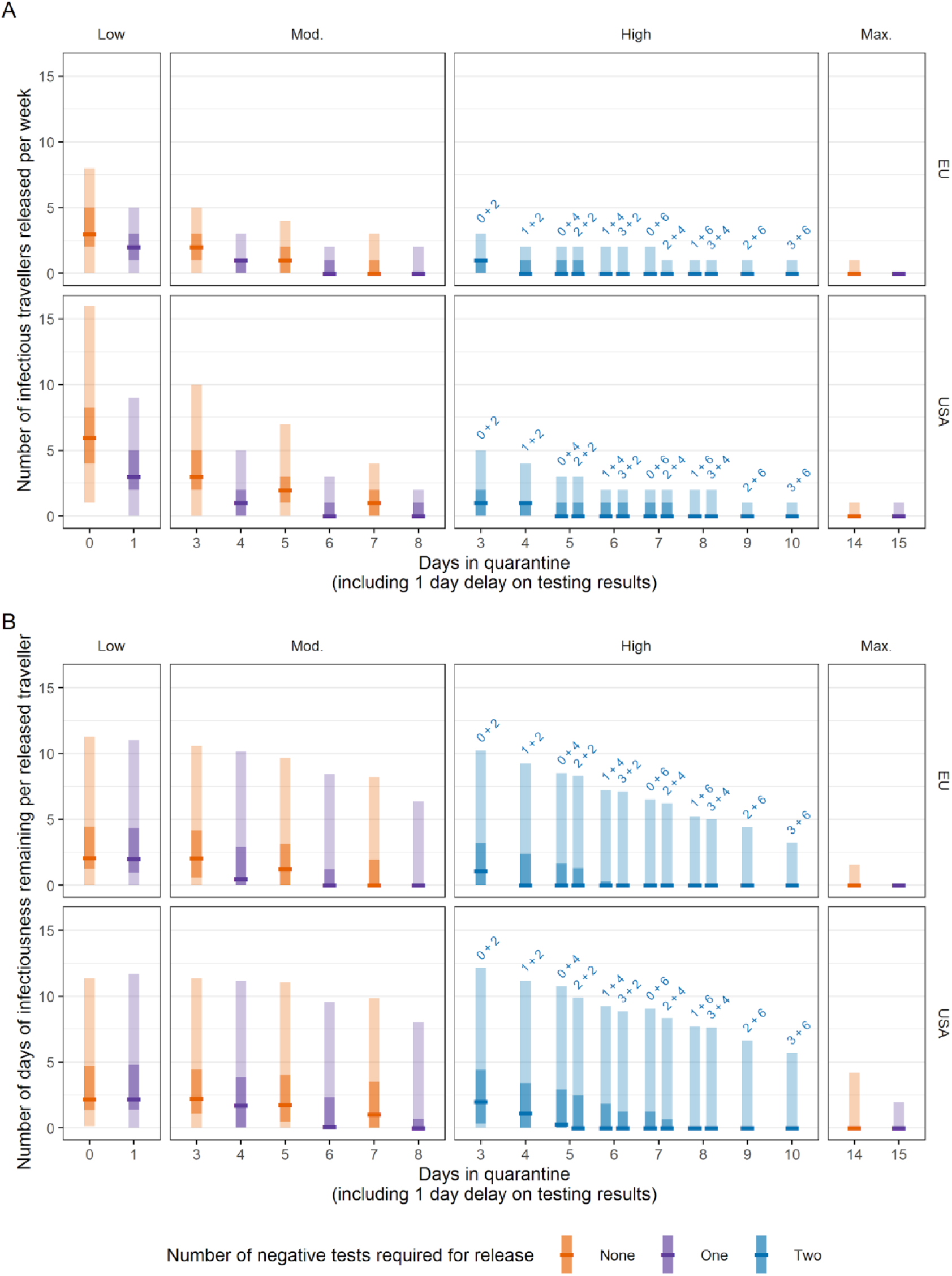
A. Expected number of infectious and pre-infectious persons free to enter the UK from the EU and USA based on observed travel volumes in each of the scenarios and how long they spend in quarantine before release, with no pre-flight testing. B. Per-individual days of infectiousness remaining after release, based on observed travel volumes. Scenarios with no testing are denoted by orange bars; single tests with purple bars, and two tests with blue bars. We assume that test results are delayed by 1 day and hence persons leave quarantine 1 day after their second test. Central bar = median; light bar = 95% uncertainty interval; dark bar = 50% uncertainty interval.

**Figure 4.**
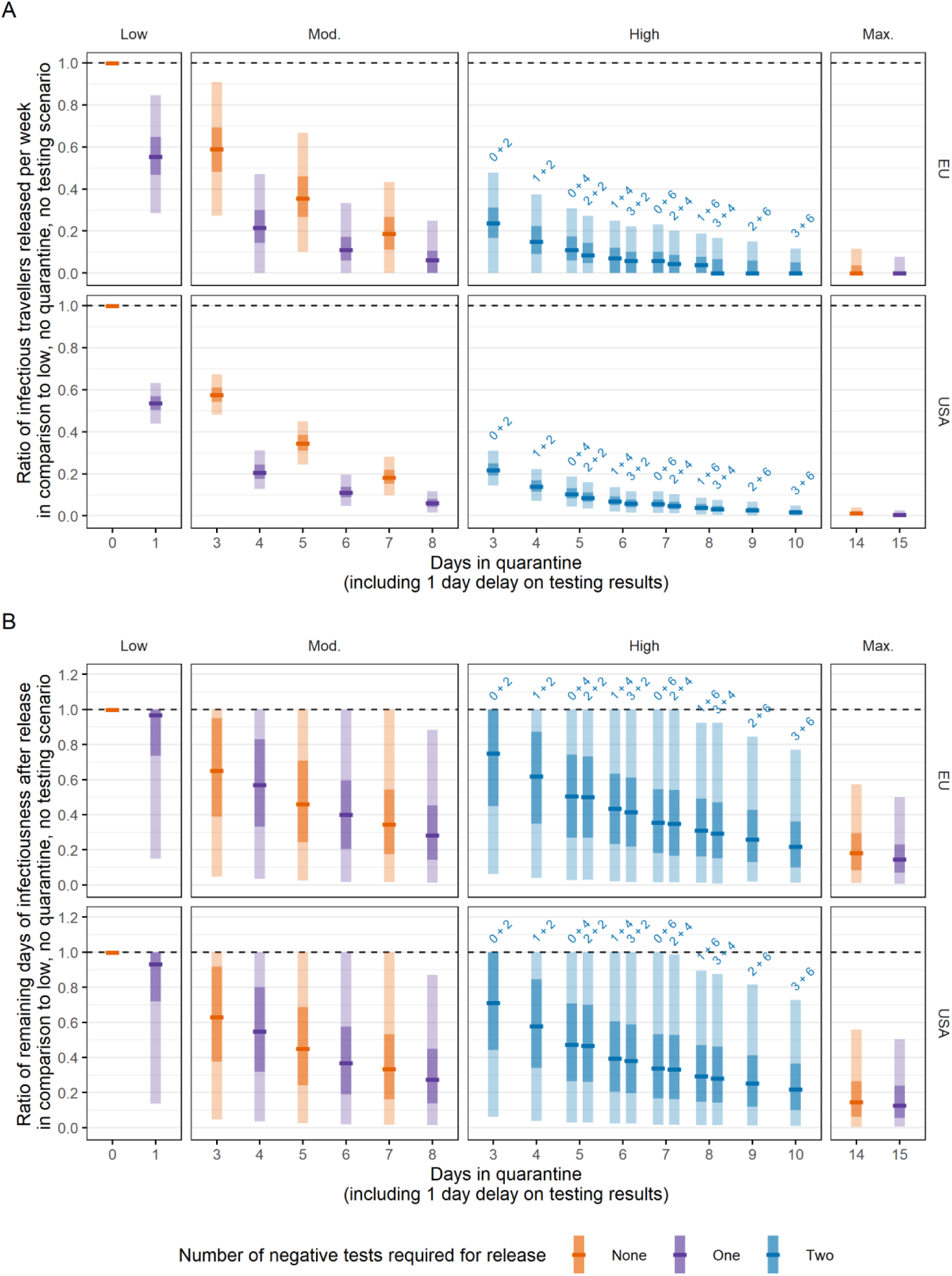
Rate ratios for the number of infectious persons released (A) and the remaining infectious person-days (B) in each scenario compared to a baseline of syndromic screening and no quarantine or PCR testing on arrival. Scenarios with no testing are denoted by orange bars; single tests with purple bars, and two tests with blue bars. We assume that test results are delayed by 1 day and hence persons leave quarantine 1 day after their second test. Central bar = median; light bar = 95% uncertainty interval; dark bar = 50% uncertainty interval.

Longer mandatory quarantine periods resulted not only in fewer infectious travellers being released into the community but also that those who were released would be at a later stage of their infectious period, with reduced potential for onward transmission. We measured this as, per-individual days of remaining infectivity (Figure 3B). Furthermore, longer quarantine periods increase the fraction of pre-symptomatic infected travellers who would have their onset of symptoms during the quarantine and hence would go into self-isolation. Accordingly, we estimated a more pronounced impact of traveller interventions on the number of infectious person-days from travellers, particularly for those who would eventually become symptomatic. The lowest stringency scenario, syndromic screening at origin and no PCR screening resulted in travellers from the EU having respectively 2.1 days (95% UI: (0, 11.2) days) of remaining infection and those from the USA having 2.2 days remaining (95% UI: (0.1, 11.3) days). Compared to only syndromic screening, a 7-day quarantine period reduced the number of remaining infectious days from travellers from the EU to a median of 0 days (mean: 1.4 days; 95% UI: (0.00, 8.7) days), a RR of 0.34 (95% UI: (0.01, 1)) and from the USA to 1.0 days (mean: 2.1 days; 95% UI: (0.00, 9.8) days), a RR of 0.33 (95% UI: (0.02, 1)). A maximally stringent 14-day quarantine reduces the median number of remaining infectious days for travellers to 0 from the EU (mean: 0.1 days; 95% UI: (0, 2.1) days) and 0 from the USA (mean: 0.3 days; 95% UI: (0, 3.6) days) (Figure 3B), with RRs of, respectively, 0.15 (95% UI: 0.01, 0.56) and 0.15 (95% UI: 0.01, 0.57) (Figure 4B).

Conducting a single test for all travellers at the end of the above quarantine periods further reduced the median number of infectious entries from the EU with RRs of 0.06 (95% UI: 0.02, 0.12) (test on day 7, release on day 8) and 0.01 (95% UI: 0.00, 0.03) (test on day 14, release on day 15) compared to the lowest stringency scenario, with similar reductions for US travellers (Figure 4A, Moderate and Max., purple). While a single test decreased the number of infectious entries, compared to quarantine alone the medians remained unchanged at 0 and the upper bounds of the 95% UI were reduced to 6.2 days (EU) and 7.7 days (USA), reflecting those still in their infectious period at the end of quarantine were more likely to be diverted to mandatory isolation.

Requiring a second round of testing in addition to quarantine and a single test had little marginal impact, although a quarantine period of over 8 days with two tests may be able to largely replicate the impact of a 14-day quarantine period (Fig 4B). For the same total time in quarantine in the High stringency scenarios, delaying the first test and reducing the time until the second test leads to the same median number of released infectious arrivals but reduces the 97.5th percentile, making the scenario more consistent. This effect is more pronounced when comparing a two-test scenario to a one-test scenario of equivalent duration (Figure S1A, Mod.).

The additional impact of pre-flight testing on the number of infectious travellers entering the community was small, and was most effective if implemented the day prior to departure in scenarios with no post-flight testing (Figure S2).

## Discussion

Here we analysed the effect of different combinations of PCR testing and self-quarantine times on the number of infectious individuals entering a country, beyond the effect of syndromic screening at departure. We find that a quarantine period of at least 5 days, with a single PCR test on the final day of quarantine, results in a reduction of over 80% in both the number of infectious persons entering the community and the transmission potential of those persons. A 7-day quarantine with a test on the final day can reduce infectious entries by an average of 94%, with little marginal benefit for additional rounds of testing. In addition, tests prior to departure appear ineffective unless conducted less than 4 days prior to departure.

Due to differences in COVID-19 prevalence and estimated travel volume in July 2020, the expected numbers of infectious entries per week in a no-intervention scenario (apart from self-reporting of symptoms and syndromic screening at departure) from the USA are approximately double that of the EU (up to 28 and up to 12, respectively). Travel volumes have been severely impacted by the pandemic, with volumes in April and May 2020 being 1% of those of 2019. We anticipate that the number of infectious travellers from each region will increasingly diverge as the number of flights and number of travellers increase, particularly as the length of quarantine precludes many travellers from taking short trips.

The risk for local COVID-19 control stemming from infectious travellers will need to be assessed in the context of the local infection incidence. For example, 6 - 11 infectious travellers per week arriving in July 2020 from the EU or USA into a community with thousands of live infections (such as that of the UK on 12 June 2020, with prevalence estimated at 45 per 10,000 (95%: (24, 92) per 10,000)) will likely have little impact on control efforts. In contrast, if local infection prevalence is lower (as in the UK on 20 July 2020: 9 (5, 18) per 10,000), these numbers of infectious travellers may pose a large risk for seeding outbreaks in the community (19). In England in late June an estimated 25,000 (13,000 to 46,000) people per week became infected with SARS-CoV-2 (37). Since then, transmission has been largely declining; however, as of mid-July 2020 still about 5,000 cases are reported per week in the UK and likely only form a small fraction of all infections in the country (16).

We presented the risk from infected travellers as the number of infectious travellers entering the community. To account for the differential residual duration on their infectiousness under the different strategies, we also presented the number of infectious person-days in the community from travellers. While the latter measure indicates an increased effectiveness of longer quarantine, it may still present an underestimate of the true effect as the measure only considers that travellers are still infectious, not how likely transmission is given their viral load declines towards the end of their infectious period (and it is likely that infectiousness declines along with it). Hence, with a peak infectivity around the onset of symptoms at about 5 days, a minimum quarantine period of 7 days is likely to release not just *fewer* infectious travellers but *less infectious* travellers, with or without the use of a test at 7 days.

Multiple assumptions were made in this analysis. We assumed that inbound travel volumes were 50% of total traveller movements as reported by the CAA, which may not reflect asymmetric patterns of repatriation. The total number of traveller movements between the UK and each of the USA and EU in April and May 2020 were approximately 1% of that reported in 2019, indicating that the combination of travel restrictions and airlines suspending flights has led to a sharp decrease in the number of potentially infected travellers. It is likely that as the pandemic progresses, travel volumes will begin to return to pre-pandemic levels and, as such, the likely number of infectious arrivals will increase unless prevalence is severely reduced. The expectation of quarantine on arrival will likely have played a key part in this reduction. In our analysis, we considered a constant air passenger volume and did not consider that shortening of quarantine will not only increase the risk from infectious travellers to the local community (19), but also increase the number of travellers. Therefore our estimates may be interpreted as change in the per traveller risk and may be underestimating the risk from an easing of the quarantine period requirements. To address this, we have provided estimates in terms of the number of infectious entries per 10,000 arrivals for the given prevalence.

Increases in prevalence at travel origins is likely to result in an increase in the expected number of infectious arrivals. Reporting delays in detecting such increases in COVID-19 risk will be made augmented by delays in amended guidelines accordingly, a process which itself is not instantaneous upon receipt of updated prevalence estimates. If prevalence is seen to be rising in a certain country, additional protection may be provided by requiring two tests to be conducted, rather than a single test, in addition to quarantine. We assumed that 70% of ever-symptomatic persons currently symptomatic (e.g with a cough or fever) would be detected or self-report and hence not travel. While COVID-19 risk awareness is now generally high, the non-specificity of symptoms will likely lead some travellers to try to ignore or suppress their symptoms and pose additional risk that we have not explicitly included in our analysis. Asymptomatic infections, who may represent a substantial proportion of all infected individuals in some locations (38), will not be prevented from boarding or have their infectious period censored by self-isolation following symptom onset. Instead, these infections can only be detected by PCR screening for ongoing infection.

We assume that the probability of detecting an asymptomatic infection is almost 40% less than that for a symptomatic infection (29). We have assumed that onset of symptoms (at peak PCR sensitivity, Figure 2A) occurs soon after the onset of infectiousness. At the median end of symptomatic cases’ infectious period, the sensitivity of PCR, when detecting symptomatic infections, is still greater than 0.6 (at the 95th percentile it is still greater than 0.4), reflecting that ever-symptomatic cases are still more likely than not to be PCR positive as well as symptomatic, and therefore spend more time than is necessary in isolation, even though they may no longer be shedding virus (29). In the high scenarios, we allow the timing of the first post-arrival test to vary between 0 days (i.e, on arrival) and 3 days (i.e when in quarantine). Requiring the first test to be administered at the airport may reduce loss to follow-up but comes at the expense of additional burden on travellers on the day of travel, extending the duration of time spent in the airport terminal.

We consider only the infectiousness of the infectious arrivals, and the ability for screening to detect them. We do not make any assumptions about the potential for self-isolating infectious travellers to infect their household upon arrival and the resulting onwards transmission, or that adherence to self-isolation may be imperfect as both a first negative test and a long duration of quarantine may reduce adherence to quarantine rules (3,5). Hence, by assuming perfect adherence, we may overestimate the added benefit of long periods of quarantine in terms of the person-days of infectiousness in the community. A policy of managed quarantine in a community facility or hotel (39) has been adopted by countries such as New Zealand and Vietnam (40) who have successfully pursued elimination strategies (41).

We assume that individuals who become symptomatic during quarantine begin an additional period of self-isolation of at least 7 days from symptom onset, until they no longer experience symptoms and their total quarantine and isolation period is at least 14 days long. The longer the duration of quarantine, the more likely it is that ever-symptomatic individuals develop symptoms and self-isolate, further reducing the number of infectious entries. This leads to the main impact of quarantine and PCR testing strategies being the reduction of possible transmission from asymptomatic travellers (Figure S3) who are only detectable here by PCR. We also assume that if individuals subsequently become symptomatic after quarantine (whether that is a result of an undetected imported SARS-CoV-2 infection with a long latent period, an infection acquired locally or abroad, or for other reasons), they follow national guidelines to immediately re-enter quarantine and seek another test as part of the local test and trace strategy. This may overestimate the ability of individuals to accurately self-diagnose their symptoms, but we assume that traveller sensitisation (42,43), particularly during quarantine, is high at this point in the pandemic.

For the foreseeable future we will need to carefully balance the need for traveller targeted intervention that reduces the likelihood of seeding local COVID-19 outbreaks with the restrictions they impose on travellers. While the acceptable number of infected travellers entering the community will depend on the local context of SARS-CoV-2 transmission we find that for travellers arriving from low prevalence destinations the absolute risk of seeding new outbreaks is likely low and hence either testing and/or quarantine-based strategies may do little to further reduce such risk, particularly when many infectious arrivals are asymptomatic cases. For arrivals from countries with ongoing community transmission, quarantine on arrival will limit the risk for onward transmission into the local community. While a 14-day quarantine will likely prevent most transmission from travellers, an 8-day quarantine (with testing on day 7) can capture as many infectious persons in approximately half the time. Testing passengers is resource-intensive but presents a way to either further reduce risks or allow a shorter quarantine at the same level of risk, particularly for arrival from countries with widespread SARS-CoV-2 transmission.

## Data Availability

All model code is available on the project Github repository.

https://github.com/cmmid/travel_screening_strategies

## Acknowledgements

The following authors were part of the Centre for Mathematical Modelling of Infectious Disease 2019-nCoV working group. Each contributed in processing, cleaning and interpretation of data, interpreted findings, contributed to the manuscript, and approved the work for publication: Katharine Sherratt, Stéphane Hué, Matthew Quaife, Nikos I Bosse, Graham Medley, Megan Auzenbergs, Adam J Kucharski, Nicholas G. Davies, Oliver Brady, Sophie R Meakin, Rein M G J Houben, Katherine E. Atkins, Kiesha Prem, C Julian Villabona-Arenas, Hamish P Gibbs, Thibaut Jombart, Charlie Diamond, Petra Klepac, Arminder K Deol, Rachel Lowe, James W Rudge, Mark Jit, Sebastian Funk, Gwenan M Knight, Simon R Procter, David Simons, Quentin J Leclerc, James D Munday, Amy Gimma, Georgia R Gore-Langton, Christopher I Jarvis, Jon C Emery, Anna M Foss, Kathleen O’Reilly, Joel Hellewell, Emily S Nightingale, Kevin van Zandvoort, Damien C Tully, Sam Abbott, Kaja Abbas, Fiona Yueqian Sun, Alicia Rosello.

## Authors’ Contributions

SC, BJQ, SFlasche and WJE conceived the study and wrote the report. SC, BJQ, AE, SFlasche and WJE designed the model. SC and BJQ led the development and analysis of the screening model and produced the results and figures. TWR led the development of and analysis of the prevalence model. TWR, YL, YWDC, CABP, AE, SFlasche, REM and WJE consulted on the analyses. The CMMID COVID-19 working group members contributed by interpreting the study findings, contributing to the report, and approving the work for publication. All authors approved the final version for publication.

We thank Anna Foss, Nikos I Bosse, James Munday, and C Julian Villabona-Arenas for their comments on a draft manuscript.

## Funding

The following funding sources are acknowledged as providing funding for the named authors. This research was partly funded by the Bill & Melinda Gates Foundation (INV-003174: YL; NTD Modelling Consortium OPP1184344: CABP). DFID/Wellcome Trust (Epidemic Preparedness Coronavirus research programme 221303/Z/20/Z: CABP). This project has received funding from the European Union’s Horizon 2020 research and innovation programme - project EpiPose (101003688: WJE, YL). HDR UK (MR/S003975/1: RME). This research was partly funded by the National Institute for Health Research (NIHR) using UK aid from the UK Government to support global health research. The views expressed in this publication are those of the author(s) and not necessarily those of the NIHR or the UK Department of Health and Social Care (16/136/46: BJQ; 16/137/109: BJQ, YL; PR-OD-1017-20002: WJE). UK MRC (MC_PC_19065 - Covid 19: Understanding the dynamics and drivers of the COVID-19 epidemic using real-time outbreak analytics: RME, SC, WJE, YL). Wellcome Trust (206250/Z/17/Z: TWR; 208812/Z/17/Z: SC, SFlasche). Alan Turing Institute and Nakajima Foundation (AE). No funding (YWDC).

The following funding sources are acknowledged as providing funding for the working group authors. BBSRC LIDP (BB/M009513/1: DS). This research was partly funded by the Bill & Melinda Gates Foundation (INV-001754: MQ; INV-003174: KP, MJ; NTD Modelling Consortium OPP1184344: GM; OPP1180644: SRP; OPP1183986: ESN; OPP1191821: KO’R, MA). BMGF (OPP1157270: KA). DFID/Wellcome Trust (Epidemic Preparedness Coronavirus research programme 221303/Z/20/Z: KvZ). DTRA (HDTRA1-18-1-0051: JWR). Elrha R2HC/UK DFID/Wellcome Trust/This research was partly funded by the National Institute for Health Research (NIHR) using UK aid from the UK Government to support global health research. The views expressed in this publication are those of the author(s) and not necessarily those of the NIHR or the UK Department of Health and Social Care (KvZ). ERC Starting Grant (#757688: CJVA, KEA; #757699: JCE, MQ, RMGJH). This project has received funding from the European Union’s Horizon 2020 research and innovation programme - project EpiPose (101003688: KP, MJ, PK). This research was partly funded by the Global Challenges Research Fund (GCRF) project ‘RECAP’ managed through RCUK and ESRC (ES/P010873/1: AG, CIJ, TJ). NIHR (16/137/109: CD, FYS, MJ; Health Protection Research Unit for Immunisation NIHR200929: NGD; Health Protection Research Unit for Modelling Methodology HPRU-2012-10096: TJ; NIHR200929: MJ; PR-OD-1017-20002: AR). Royal Society (Dorothy Hodgkin Fellowship: RL; RP\EA\180004: PK). UK DHSC/UK Aid/NIHR (ITCRZ 03010: HPG). UK MRC (LID DTP MR/N013638/1: GRGL, QJL; MC_PC_19065 - Covid 19: Understanding the dynamics and drivers of the COVID-19 epidemic using real-time outbreak analytics: AG, NGD, TJ; MR/P014658/1: GMK). Authors of this research receive funding from UK Public Health Rapid Support Team funded by the United Kingdom Department of Health and Social Care (TJ). Wellcome Trust (206250/Z/17/Z: AJK; 206471/Z/17/Z: OJB; 210758/Z/18/Z: JDM, JH, KS, NIB, SA, SFunk, SRM). No funding (AKD, AMF, DCT, SH).

## Conflicts of interest

Akira Endo received a research grant from Taisho Pharmaceutical Co. Ltd.

## Supplementary appendix

### Code and data sharing

The model code is available at https://github.com/cmmid/travel_screening_strategies.

**Figure S1.**
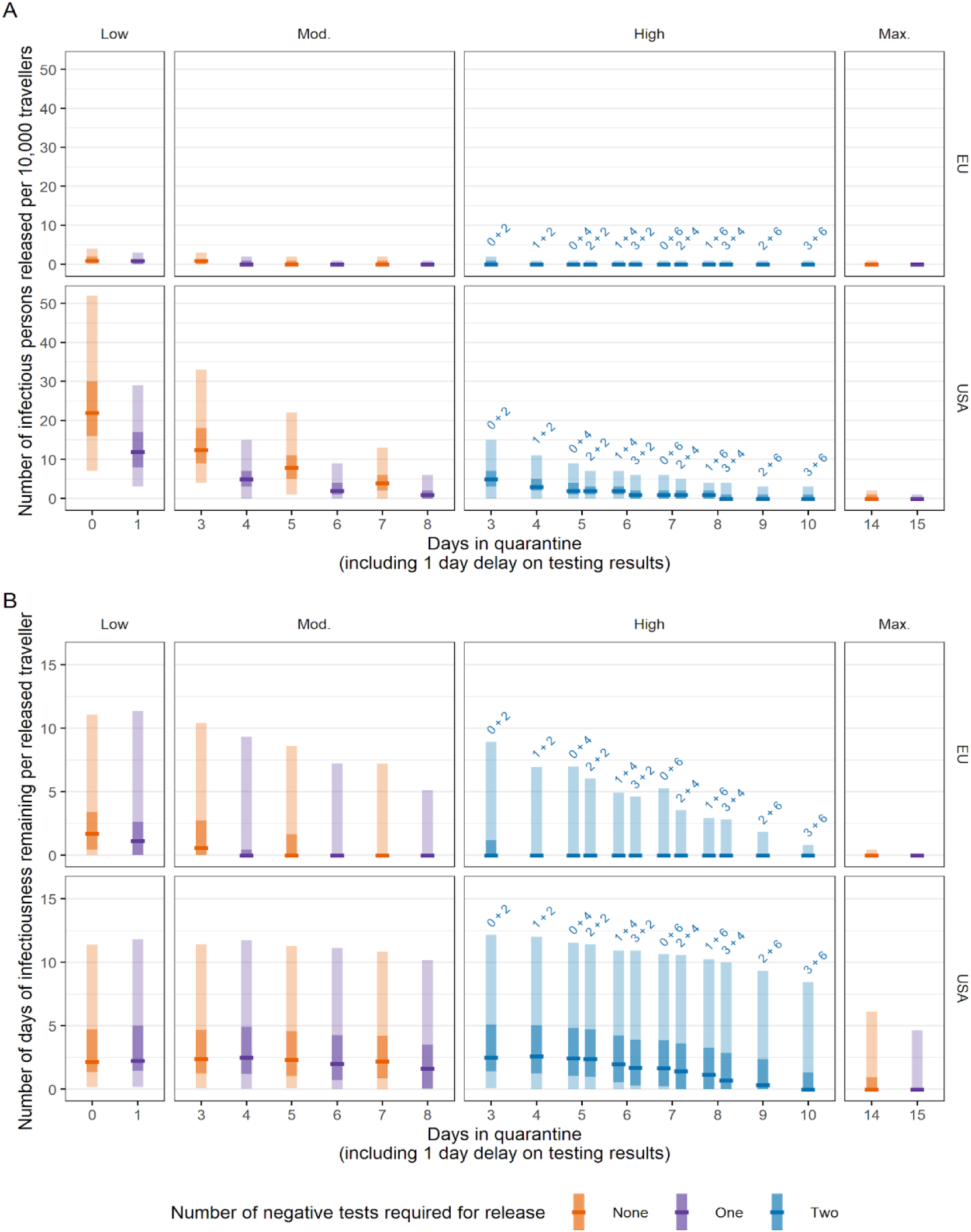
A. Expected number of infectious and pre-infectious persons per 10,000 travellers free to enter the UK from the EU and USA in each of the scenarios and how long they spend in quarantine before release, with no pre-flight testing. B. Per-individual person-days of infectiousness remaining after release per 10,000 travellers. Persons showing symptoms at departure were assumed to be prevented from travel, and post-infectious persons were assumed to not carry any risk of seeding transmission. Scenarios with no testing are denoted by orange bars; single tests with purple bars, and two tests with blue bars. We assume that test results are delayed by 1 day and hence persons leave self-isolation 1 day after their second test. Central bar = median; light bar = 95% uncertainty interval; dark bar = 50% uncertainty interval.

**Figure S2.**
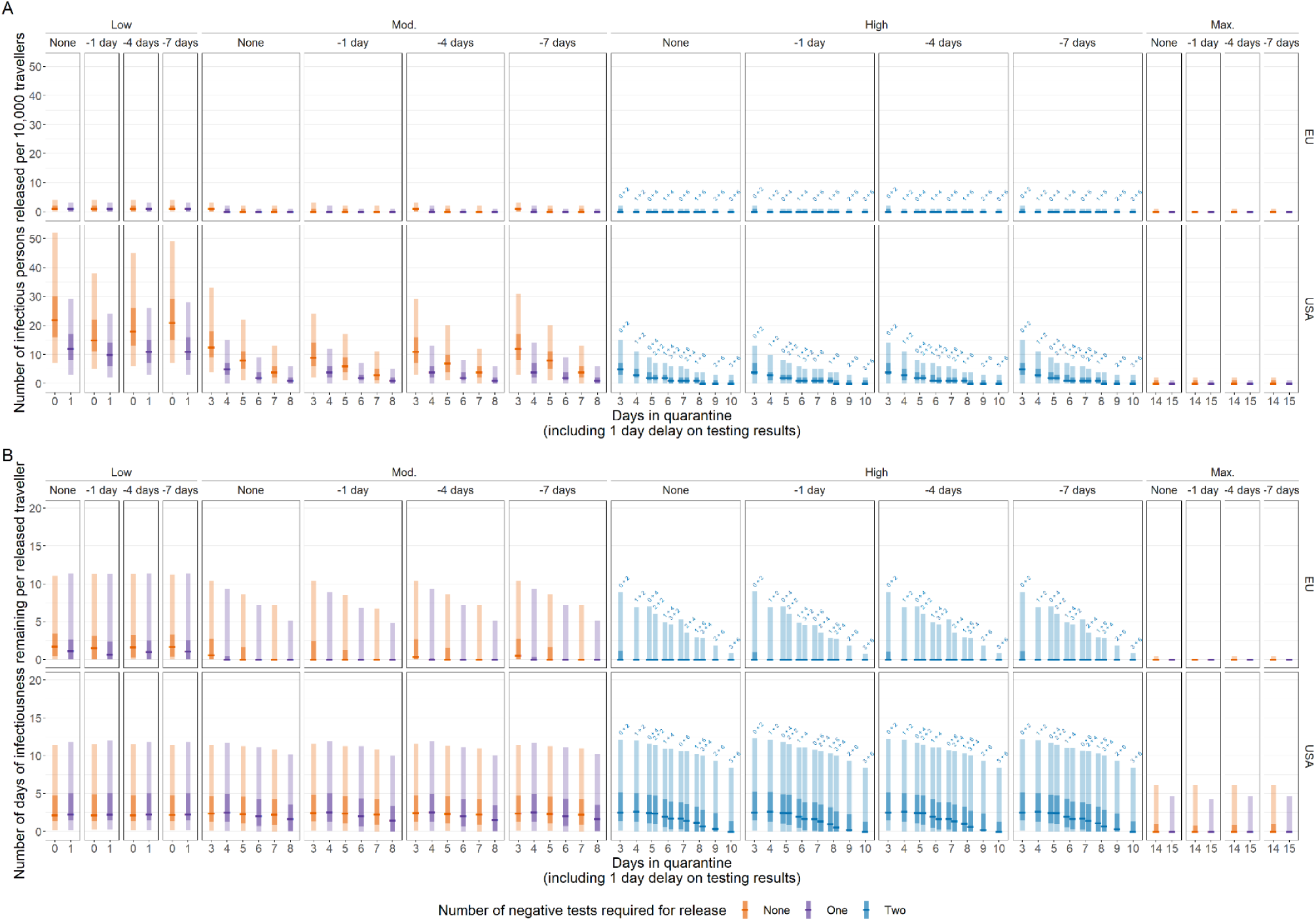
No pre-flight testing or pre-flight testing 1, 4 or 7 days prior to the flight in addition to each of the scenarios. Expected number of infectious and pre-infectious persons free to enter the UK from the EU (top) and USA (bottom) per 10,000 travellers in each of the scenarios and how long they spend in isolation before release. Persons showing symptoms at departure were assumed to be prevented from travel, and post-infectious persons were assumed to not carry any risk of seeding transmission. Scenarios with no testing are denoted by orange bars; single tests with purple bars, and two tests with blue bars. We assume that test results are delayed by 1 day and hence persons leave self-isolation 1 day after their second test. Central bar = median; light bar = 95% uncertainty interval; dark bar = 50% uncertainty interval.

**Figure S3.**
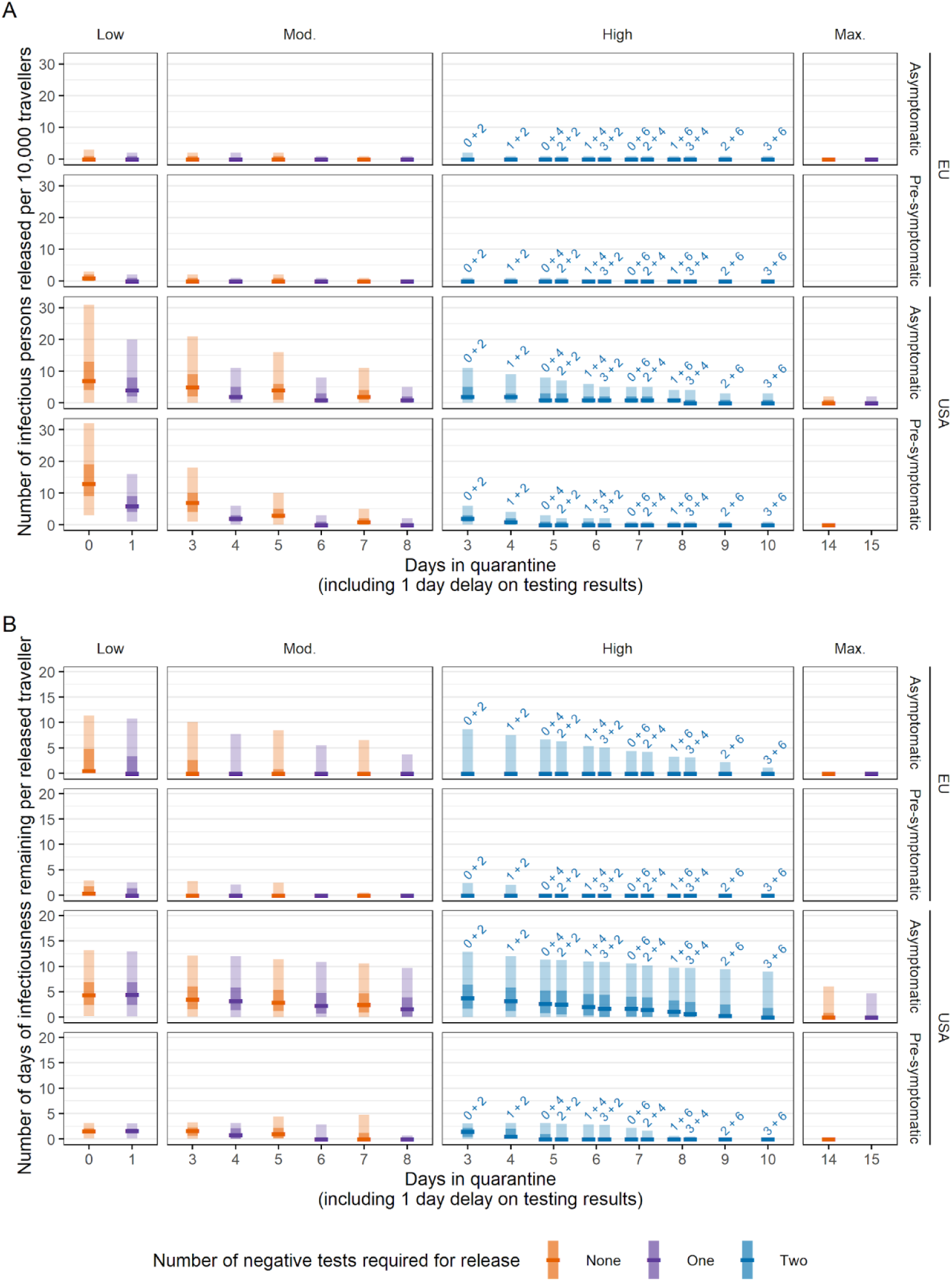
Stratified on asymptomatic or pre-symptomatic. A. Expected number of infectious and pre-infectious persons per 10,000 travellers free to enter the UK from the EU and USA in each of the scenarios and how long they spend in quarantine before release, with no pre-flight testing. B. Cumulative person-days of infectiousness remaining after release per 10,000 travellers. Persons showing symptoms at departure were assumed to be prevented from travel, and post-infectious persons were assumed to not carry any risk of seeding transmission. Scenarios with no testing are denoted by orange bars; single tests with purple bars, and two tests with blue bars. We assume that test results are delayed by 1 day and hence persons leave quarantine 1 day after their second test. Central bar = median; light bar = 95% uncertainty interval; dark bar = 50% uncertainty interval.

**Figure S4.**
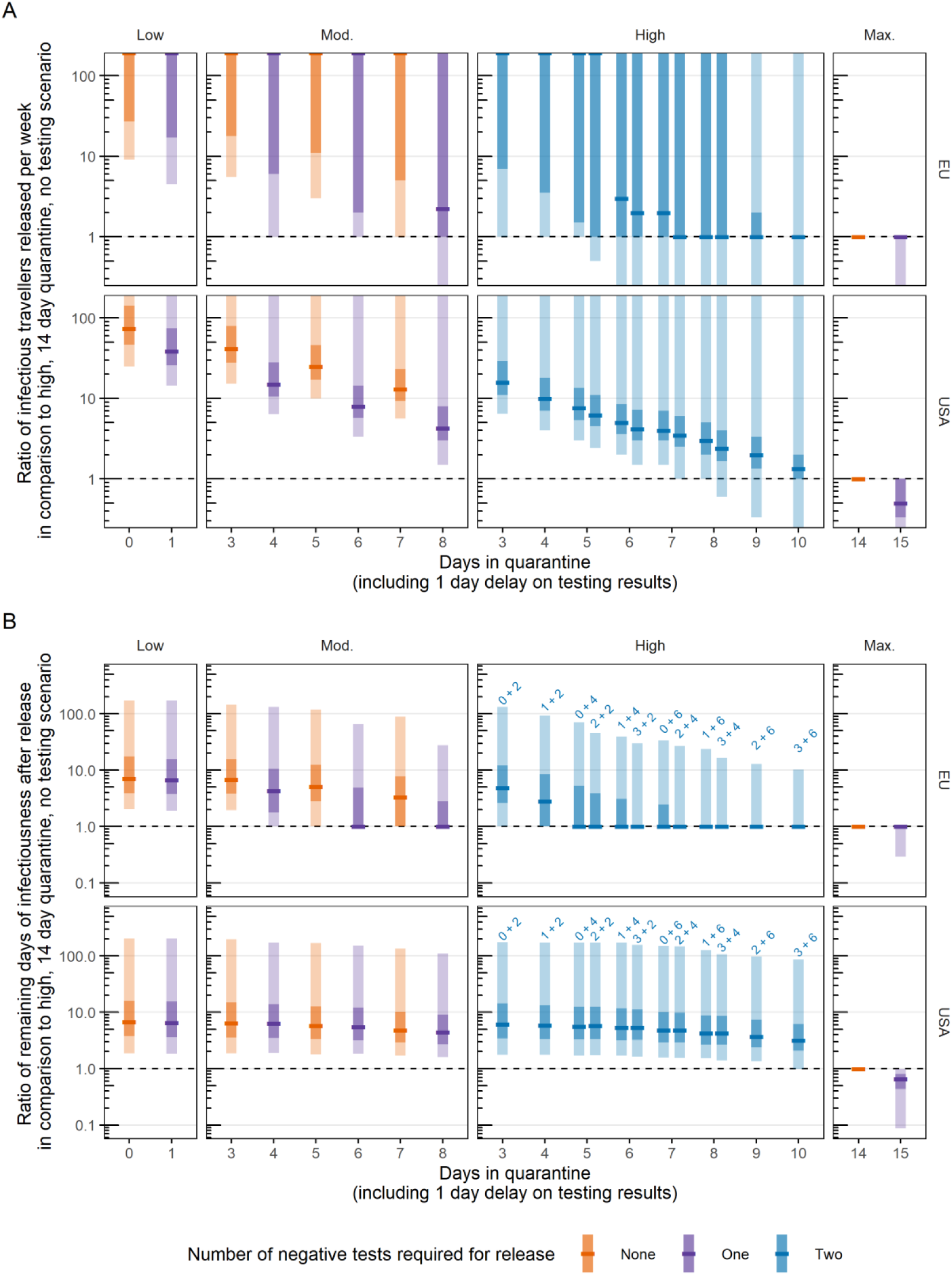
Rate ratios for the number of infectious persons released (A) and the remaining infectious person-days (BA) in each scenario compared to a baseline of a 14 day mandatory quarantine and no PCR testing on arrival. Scenarios with no testing are denoted by orange bars; single tests with purple bars, and two tests with blue bars. We assume that test results are delayed by 1 day and hence persons leave quarantine 1 day after their second test. Central bar = median; light bar = 95% uncertainty interval; dark bar = 50% uncertainty interval. Where the median is at the upper limit of the plot, more than 50% of simulations have an infinitely large ratio.

**Figure S5.**
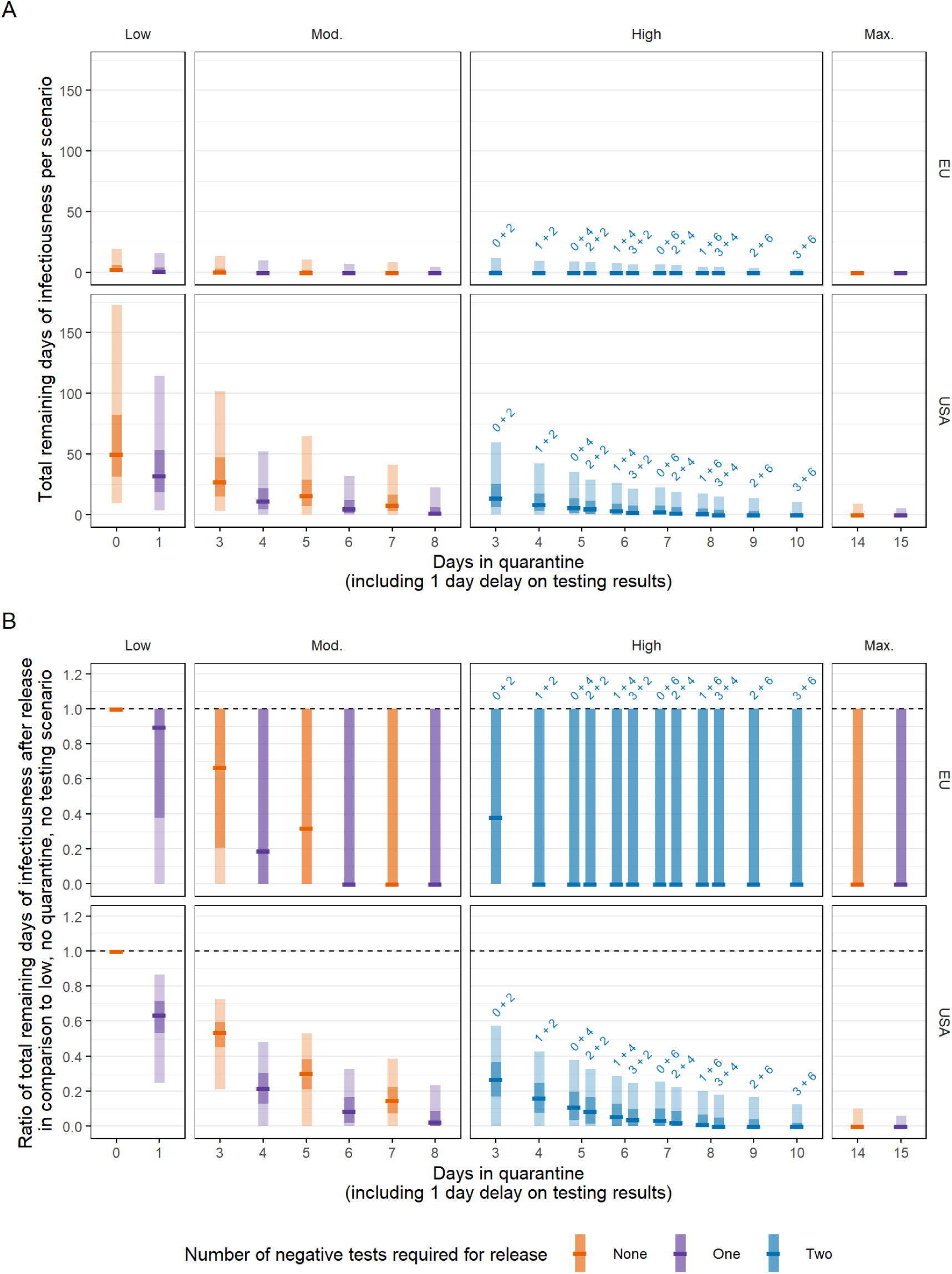
A) Cumulative person-days of infectiousness remaining in each scenario with 10,000 travellers. B) Ratio of cumulative person-days remaining compared to a baseline of syndromic screening and no quarantine or PCR testing on arrival. Scenarios with no testing are denoted by orange bars; single tests with purple bars, and two tests with blue bars. We assume that test results are delayed by 1 day and hence persons leave quarantine 1 day after their second test. Central bar = median; light bar = 95% uncertainty interval; dark bar = 50% uncertainty interval.

